# The seasonality of three childhood infections in a pre-industrial society without schools

**DOI:** 10.1101/2021.10.08.21264734

**Authors:** Michael Briga, Susanna Ukonaho, Jenni E Pettay, Robert J Taylor, Tarmo Ketola, Virpi Lummaa

**Affiliations:** Department of Biology, University of Turku, Vesilinnantie 5, 20014 Turku, Finland; Infectious Disease Epidemiology Group, Max Planck Institute for Infection Biology, Charitéplatz 1, Campus Charité Mitte, 10117 Berlin, Germany; Department of Biological and Environmental Science, University of Jyväskylä, P.O. Box 35, FI-40014 Jyväskylä, Finland; Department of Science and Environment, Roskilde University, 4000 Roskilde, Denmark

## Abstract

**Background:** The burden of many infectious diseases varies seasonally and a better understanding of the drivers of infectious disease seasonality would help to improve public health interventions. For directly transmitted highly-immunizing childhood infections, the leading hypothesis is that seasonality is strongly driven by social gatherings imposed by schools, with maxima and minima during school terms and holidays respectively. However, we currently have a poor understanding of the seasonality of childhood infections in societies without schools and whether these are driven by human social gatherings. Here, we used unique nationwide data consisting of >40 epidemics over 100 years in 18^th^ and 19^th^ century Finland, an agricultural pre-health care society without schools, to (i) quantify the seasonality of three easily identifiable childhood infections, smallpox, pertussis and measles and (ii) test the extent to which seasonality of these diseases is driven by seasonal social gatherings.

**Methods:** We quantified the seasonality of transmission using time series Susceptible-Infected-Recovery models, wavelet analyses and general additive mixed models.

**Results:** We found that all three infections were seasonal and the seasonality patterns differed from those in industrialized societies with schools. Smallpox and measles showed high transmission in the first half of the year, but we could not associate this with seasonal human gatherings events. For pertussis, however, transmission was higher during social gathering events such as New Year and Easter.

**Conclusions:** Our results show that the seasonality of childhood infections is more variable than previously described in other populations and indicate a pathogen-specific role of human social aggregation in driving the infectious disease dynamics.

**Funding:** Academy of Finland (278751, 292368), Nordforsk (104910), the Ehrnrooth Foundation, the Finnish Cultural Foundation, the University of Turku Foundation and the Doctoral Programme in Biology, Geography and Geology, University of Turku.

**Clinical trial number:** NA

## 1. Introduction

Many infectious diseases are seasonal, in that they tend mostly to circulate during only part of the year (Altizer et al. 2006; Grassly and Fraser 2006; Bakker et al. 2016; Martinez 2018). Quantifying this seasonality and identifying its drivers is of interest for our fundamental understanding of the spatio-temporal dynamics of infectious diseases (Earn et al. 2000; Rohani et al. 2002; Ferrari et al. 2008) and for public health authorities to develop appropriate intervention measures (Stone et al. 2007; Cauchemez et al. 2009).

In industrialized societies, the leading explanation for the seasonality of many infections such as measles and influenza is a phenomenon called “school-term forcing,” in which social gatherings associated with the school term increase transmission (London and Yorke 1973; Fine and Clarkson 1982; Deguen et al. 2000; Bjørnstad et al. 2002; Cauchemez et al. 2008; Eames et al. 2012). However, the vast majority of studies on the seasonality of childhood infections were carried out in societies with schools. This can be a limitation because, even though contact is a necessary condition for transmission, the bias towards societies with schools can obscure co-variation with other seasonal factors such as climate, other circulating infections and the host’s immune system (Metcalf et al. 2017). For example, in many industrialized countries, schools are on holiday between June and September, which are often the hottest and driest months thereby limiting the transmission of many droplet-mediated childhood infections such as measles and pertussis.

A useful approach to identify the role of seasonal social gathering is to compare the seasonality of infections before and after the start of systematic schooling, but pre-school data remain rare, thereby preventing such comparisons, and our understanding of the seasonality of childhood infections in pre-industrial societies remains poor (Duncan et al. 1996, 1997; Conlan and Grenfell 2007; Krylova and Earn 2020). Here we report an extensive analysis of a unique nationwide dataset of numbers of deaths from three childhood infections, smallpox, pertussis and measles (Table 1), occurring over a span more than 100 years in more than 200 municipalities (parishes) in 18^th^ and 19^th^ century agricultural Finland. At that time, Finland’s educational system was characterized by traveling schools (kiertokoulu), whereby an educator would visit a parish for a few weeks and then travel to a next parish (Tiimonen 2001) and there was essentially no health care (Saarivirta et al. 2012), which allows for a natural spread of infections.

**Table 1.** Overview of the infections in this study, with their sample sizes before and after the start of the smallpox vaccination program, respectively the pre-vaccine era (1750-1801) and vaccine era (1802-1850).

| **Infection** | **Smallpox** |  | **Pertussis** |  | **Measles** |  |
| --- | --- | --- | --- | --- | --- | --- |
| pathogen | virus |  | bacteria |  | virus |  |
| generation time | 2 weeks |  | 4 weeks |  | 2 weeks |  |
| immunization | long-lasting |  | waning |  | long-lasting |  |
| vaccination | vaccine |  | none |  | none |  |
| Sample size | 1750-1801 | 1802-1850 | 1750-1801 | 1802-1850 | 1750-1801 | 1802-1850 |
| N deaths | 39,082 | 30,629 | 14,385 | 31,175 | 7,429 | 15,153 |
| N deaths/year | 752 | 638 | 277 | 649 | 143 | 316 |

Our first aim was to characterize the seasonality of three childhood infections smallpox, pertussis and measles in this historical pre-industrial society without schools. Because education did not have a clear seasonality, we hypothesized that the seasonality of childhood infections would be different from that of school-term societies in contemporary industrialized countries. However, pre-industrial Finland did have seasonal social gathering events such as New Year, Easter and solstice celebrations, and sowing and harvest gatherings (Vilkuna 1950; Karjalainen 1994). Hence, our second aim was to test the hypothesis that these seasonal social gathering events drove the observed seasonality of childhood infections (Guyer and Mcbean 1981; Ferrari et al. 2010). To this end, we used time-series Susceptible-Infected-Recovery (tSIR) models to estimate pathogen transmission rates and then assessed statistically whether weeks with seasonal social gathering events were associated with higher pathogen transmission.

## 2. Materials and Methods

### 2.1 Study population and data

Our data on births, deaths and causes of deaths across Finland were originally recorded by Lutheran priests by the order of the King, and subsequently digitized by the Genealogical Society of Finland HisKi project, and are available at: http://hiski.genealogia.fi/historia/indexe.htm. The database consists of 5,884,901 individual births and 3,490,737 individual deaths occurring between 1600 and 1948 in 507 parishes. For the purpose of the analyses here we focused on the years with most complete birth and causes of death data, i.e. between 1750 and 1850. This period can be characterized as pre-industrial, with primitive agricultural technology and limited health care, high birth and death rates (Holopainen and Helama 2009; Saarivirta et al. 2012) and an increase in population size from 425,700 to 1,628,900 individuals (Official Statistics of Finland 2018).

We selected those parishes without missing years in the data and below the Arctic circle (66°33′47.1’’). The parish areas above the Arctic Circle were different in many ways, with smaller population sizes occupying vast geographic areas mostly consisting of Saami, a nomadic population of reindeer herders who depended on fishing and hunting for their livelihood with little or no agriculture (Itkonen 1948). Based on these criteria, we used data from 215 parishes, located in‘southern’ Finland containing 1,693,056 birth and 1,193,072 death records, which covered 50% (interannual SD: 5%) of the Finnish population recorded in this period (Pitkänen 2007; Official Statistics of Finland 2018; Voutilainen et al. 2020). For more information on these parishes and their mortality due to childhood infections see Ketola et al. (2021).

In the historical parish records, identification of the death causes was done by the parish priests. The historical records contain 51,075 causes of death, recorded in Swedish, Finnish or German. Identification of the causes of interest was first done by classifying causes according to their spelling similarity, their synonyms in different languages and following (Vuorinen 1999). This was done by two investigators independently (M.B and T.K.) and resulted in a consistent outcome with the same causes of death being attributed to smallpox, pertussis and measles. Using this protocol, we identified 70,704 deaths due to smallpox, 46,259 due to pertussis and 22,646 due to measles and these are the data we used in the manuscript (Table 1, Fig. 1).

**Fig. 1.**
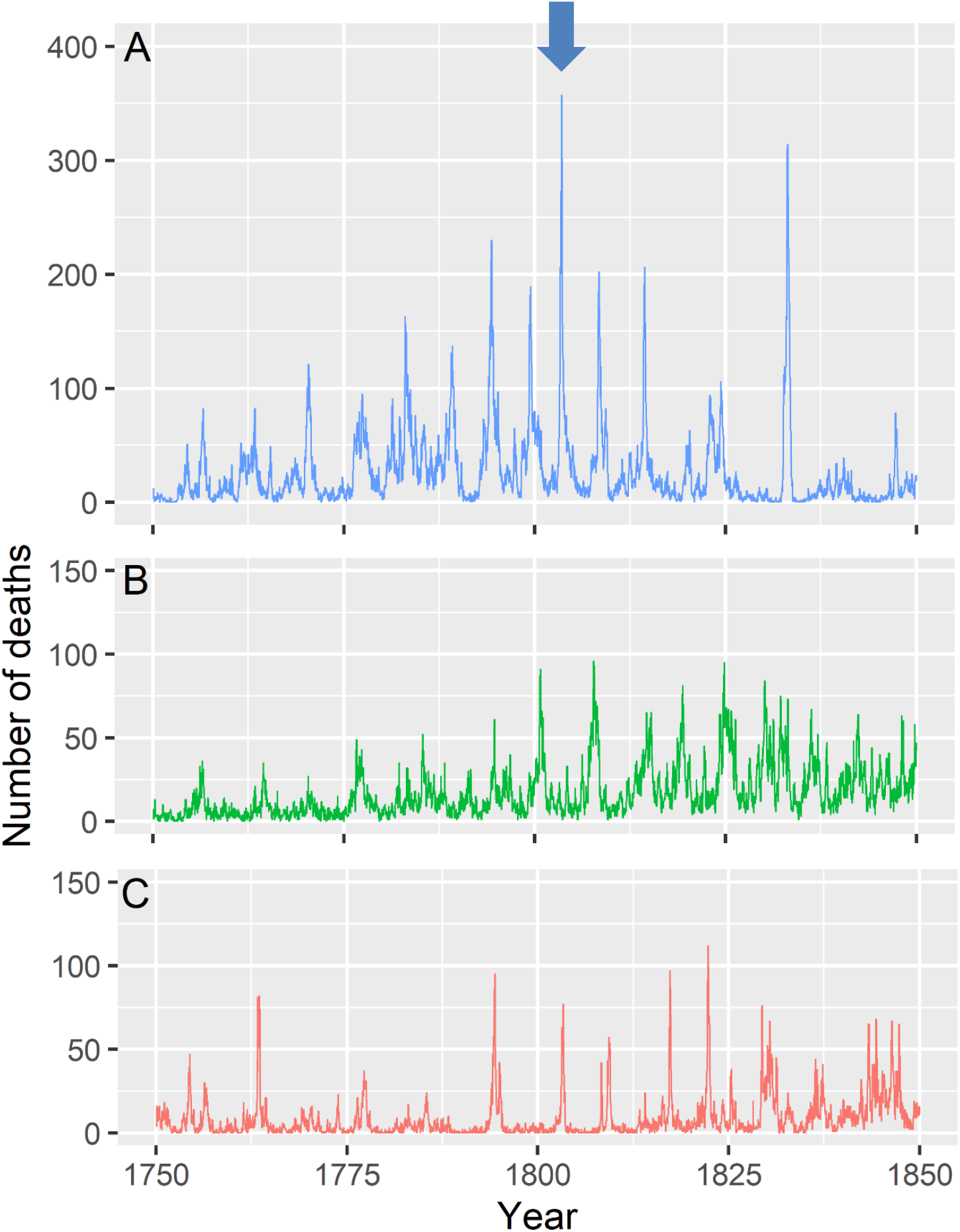
Bi-weekly counts smallpox, pertussis and measles deaths showing over 40 recurrent epidemics of between 1750 and 1850 in Finland. Blue arrow indicates the start of the smallpox vaccination programme in Finland in 1802, but for the other infections there was no vaccine available at the time. Note the different Y-axes between A and B or C.

We chose three infections: the viral infections smallpox and measles and the bacterial infection pertussis (Table 1; Anderson and May 1991). We chose these three infections for five reasons. First, the identification of the death causes was done by the parish priests and smallpox, pertussis and measles are straightforward to diagnose: smallpox and measles have characteristic rashes and pertussis has a distinct cough. Second, all three infections are transmitted either through direct contact or via droplets (Benenson 1981). Third, the three infections have similar generation times (∼the latent plus infectious period, Anderson and May 1991), i.e. two weeks for smallpox and measles and four weeks for pertussis. Fourth, in our population, these infections affected mostly young children also in historical Finns, with a mean age at death of 1.5 years for pertussis (95% CI: 0.5-8.5), 2.9 years for measles (95% CI: 0.5-13.5) and 3.8 years for smallpox (95% CI: 0.5-18.5). Fifth, these three infections were leading causes of childhood mortality in 18^th^ and 19^th^ century Finland (Table 1, Fig. 1).

We obtained the timing of seasonal social gathering events in 18^th^ and 19^th^ century Finland from two reference works (Vilkuna 1950; Karjalainen 1994). Because the descriptions of social gathering events are qualitative rather than quantitative, two investigators (J.P. & S.U.) independently identified the weeks during which these events occurred. The two determinations were consistent with each other, finding five periods of recurring seasonal social gatherings: (i) Christmas and New Year celebrations (weeks 1 and 52), (ii) Midwinter (laskiainen; weeks 5 to 9), (iii) the late-winter and spring gatherings of Easter, Walpurgis Night, Pentecoste and Midsummer solstice (weeks 12 to 25), (iv) the summer harvest (weeks 28 to 32) and (v) the autumn harvest (weeks 39 to 44). The seasonal social gatherings are shown as intervals because the week at which they occurred can vary between years. Similarly, for (iii) the spring gathering event, we merged Easter, Walpurgis Night, Pentecoste and Midsummer solstice into one seasonal event because their interannual variation in weeks of occurrence often overlap.

### 2.2 Data analysis

#### 2.2.1 General approach

In this study we had two aims. First, we identified whether the three childhood infections show seasonality (Aim 1). Second, we tested whether weeks with seasonal social gatherings have increased infectious disease transmission (Aim 2). To accomplish these two aims, we used three methods: (i) we estimated the seasonality of transmission rates from time-series Susceptible-Infected-Recovery (tSIR) models, (ii) to provide an independent confirmation of the tSIR results in (i) we repeated the analysis using wavelet analyses, and (iii) we tested whether weeks with seasonal social gatherings have increased infectious disease transmission using general additive models (GAMs).

#### 2.2.2 Method 1: Identifying infectious disease seasonality using tSIR-models

To estimate variation in disease transmission per era, we used the time-series susceptible-infected-recovered (tSIR) model as developed in Bjørnstad et al. (2002). To fit the tSIR model to the data, we followed previously established techniques which are implemented in R version 4.0.2 (R Core Team 2020) with the function‘estpars’ of the package‘tsiR’ (Becker and Grenfell 2017). In brief, in the tSIR model the population is divided into Susceptibles S, Infected I and Recovered R. The parameter of interest is the transmission factor β, which captures the rate at which individuals transfer from the S to I and hence transmit the infection. The time series approach divides time in discrete steps which reflect the diseases’ generation time, which is two weeks for smallpox and measles and four weeks for pertussis (Anderson and May 1991). It is possible that there is a time lag between onset of disease and death, which we did not include in the tSIR models. While having an estimate of such time lag from historical pre-industrial societies is difficult, we note that tSIR models with generation times that were two weeks longer than those presented here, a time possible lag in infants as indicated by some studies on smallpox and pertussis in 20^th^ century industrialized societies (Downie et al. 1969; Centers for Disease Control and Prevention 2002), gave results consistent with those presented here.

To estimate β, we first reconstructed the susceptible dynamics. At each time step, the number of susceptibles S_t_ fluctuates around a mean S_m_ such that S_t_ = S_m_ + Z_t_. Based on pre-vaccination data from the United Kingdom, S_m_ was fixed at 0.035 (Bjørnstad et al. 2002; Becker and Grenfell 2017). Note however that analyses with other values of S_m_ (0.015-0.15) gave conclusions consistent with those presented here (results not shown). The deviation of susceptibles Z_t+1_ at each time step is then given by:

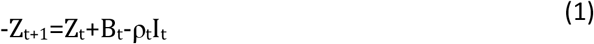

B_t_ and I_t_ are the number of births and infected respectively as given by the data. ρ_t_ is a time varying correction factor for underreporting. However, because our data reports only deaths, ρ_t_ here is a combined measure of underreporting and fatality rate. It can be estimated using the regression between cumulative births and cumulative observed cases. Given these parameters, the dynamics of the deviation susceptibles Z_t+1_ can be estimated by the residuals of the regression model in (1), which for small populations as in pre-industrial Finland, is best captured by a Gaussian process (Caudron et al. 2015).

Following the tSIR model, the infection dynamics at each time step can be described by:

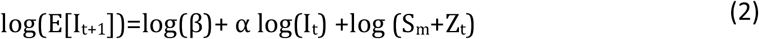

where the expected number of infected individuals E[I_t+1_] is given by the data. α is a correction factor that accounts for the discretization of a continuous time process and for inhomogeneous population mixing. We here fixed α=0.97, a common value that impacts transmission dynamics independently of epidemic size (Bjørnstad et al. 2002; Caudron et al. 2015; Becker and Grenfell 2017). Note that minor variations in α values (0.95-0.99) gave results that are consistent with those reported here. In equation (2) the only remaining unknown parameter is β, which can be estimated at each time point as the residuals of the regression of log(E[I_t+1_]) on log(I_t_) with log (S_m_+Z_t_) as an offset. For this regression, we used a generalized linear model with a Gaussian distribution and log link function (Caudron et al. 2015) but other distributions gave conclusions that are consistent with those reported here (results not shown).

#### 2.2.3 Method 2: Confirming the results of Method 1 using wavelet analyses

To provide an independent statistical test of the seasonality of infectious diseases, we performed wavelet analyses. In brief, wavelet analysis decomposes time series data using functions (wavelets) simultaneously as a function of both period and time, thereby making it possible to detect changes in periodicity, such as annual seasonality, over time (Cazelles et al. 2007). We pooled the data into monthly intervals and tested periods between six and 18 months with 12 months showing annual seasonality. To determine statistical significance, we compared the periodicity of the data with that of 1000‘white noise’ datasets with significance at the 5% level. We performed wavelet analyses following (Cazelles et al. 2007; Roesch and Schmidbauer 2018), in which we log-transformed (+1) the monthly data and fitted a Morlet wavelet with the function‘analyze.wavelet’ using the package‘WaveletComp’ (Roesch and Schmidbauer 2018).

#### 2.2.4 Method 3: Identifying drivers of infectious disease seasonality using general additive models

To statistically test whether transmission rates were higher during weeks of seasonal social gatherings events we started off with a trivariate (smallpox, pertussis, measles) general additive model (GAM) using the function‘gamm’ from the package‘mgcv’ (Woods et al. 2016; Woods 2017). We determined the‘statistical significance’ of terms and models based on (i) the model selection approach using the second order Akaike’s Information Criterion (AICc) and (ii) classic p-values. Model fitting should be considered as a continuum for which alternative models within 4 ΔAICc are plausible and become increasingly equivocal up to 14 ΔAICc, after which they become implausible (Burnham and Anderson 2002; Burnham et al. 2011). We also checked the terms p-values at p=0.05.

In our models, the dependent variable was the infection’s transmission rates estimated from the tSIR-model. To capture the interannual variance in the transmission of infection, we estimated the transmission coefficients from tSIR analyses in four-year intervals. We chose four-year intervals because it was the shortest time interval between epidemics (Fig. 1, Briga et al. unpublished), but note that analyses with any intervals up to 16 years (25% of the sample size) gave conclusions consistent with those presented here (results not shown). We also ran all analyses with four-year running means, which also gave conclusions consistent with those presented here (results not shown). To avoid that some years influenced the results more than others, we standardized all transmission rates to an annual mean of zero and, to avoid that the results be driven by a few influential some seasonal maxima, we log-transformed (+1) all transmission rates.

We had two types of predictor variables in our GAM models. First, we included‘social’, a six-level factor that reflected whether the week belonged to either one of the five aforementioned seasonal social gathering events or was‘asocial’. Second, because human seasonal social gatherings correlate with seasonal climatic cycles (e.g., agricultural activities are inevitably climate bound and social activity may be greater during summer) we tested whether the effect of social gathering events on transmission remained statistically significant when including gradual cyclic changes by adding the predictor variable‘daylength’ as a smoothed term using cubic regression splines (note that any spline gave conclusions consistent with those shown here). We obtained the daylength data from 1750 until 1850 at the location of Helsinki from the US National Oceanic and Atmospheric Administration (NOAA) daylength calculator at https://gml.noaa.gov/grad/solcalc/index.html.

We started off our GAM models with a trivariate analysis to capture a possible covariation between dependent variables. However, because multivariate GAMs do not allow inclusion of random terms or temporal autocorrelation of the time series, we performed follow-up univariate analyses including temporal autocorrelation as an auto-regressive factor of the order one (corAR1; −17.4<ΔAICc<-79.7) and year as a random intercept even though it provided a somewhat worse model fit (ΔAICc=+2 for all infections). The co-variation between the transmission rates of all three infections remained low (<0.1 in the best fitting model in Table S1) and conclusions from multivariate and univariate analyses were always consistent, though multivariate models were often more conservative than univariate models (Tables S1 & S2), and we formulated our conclusions based on the most conservative approach. We tested a variety of smoothing functions, which gave consistent results and here we present all models using cubic splines‘cs’. Model residuals fulfilled all assumptions as controlled with the functions‘resid’ and‘gam.check’ from the package‘mgcv’ (Woods et al. 2016; Woods 2017).

## 3. Results

### 3.1 Aim 1: To what extent do smallpox, pertussis and measles show seasonality?

At least a dozen epidemics of each of the three infections (smallpox, pertussis and measles) occurred between 1750 and 1850 (Table 1, Fig. 1). Because public health interventions may change the seasonality of infections, we quantified the seasonality of infections into two eras: before and after the introduction of the smallpox vaccine, respectively from 1750 until 1801 and from 1802 until 1850. Although there were no vaccines for pertussis and measles at that time, we nonetheless preferred to use the two eras for these infections as well because of possible interactions between infections (Rohani et al. 2003).

We found evidence of seasonality for all three infections in both eras (Fig. 2, Fig. S1, Fig. S2). For smallpox and measles, transmission (i.e. β in the tSIR models) was generally higher in the first half of the year than the second, in both eras (Fig. 2A & 2C, Fig. S1A & C). For smallpox in the pre-vaccine era, transmission in the first half of the year was 6% higher than the era mean, while in the vaccine era the change was larger at 19% higher than the era mean (Fig. 2A, Fig. S1A). For measles, transmission in the first half of the year was 19% and 3% higher relative to the pre-vaccine and vaccine era means, respectively (Fig. 2C, Fig. S1C). In both eras, the highest transmission occurring during week 16 with maxima at or close to 60% higher than the era means (Fig. 2C, Fig. S1C). For smallpox and measles, some of the seasonal maxima were larger than the 95% CI around the eras’ mean transmission (Fig. S1A & C), indicating statistically significant seasonality and this was confirmed through wavelet analyses (Fig. S2A & C).

**Fig. 2.**
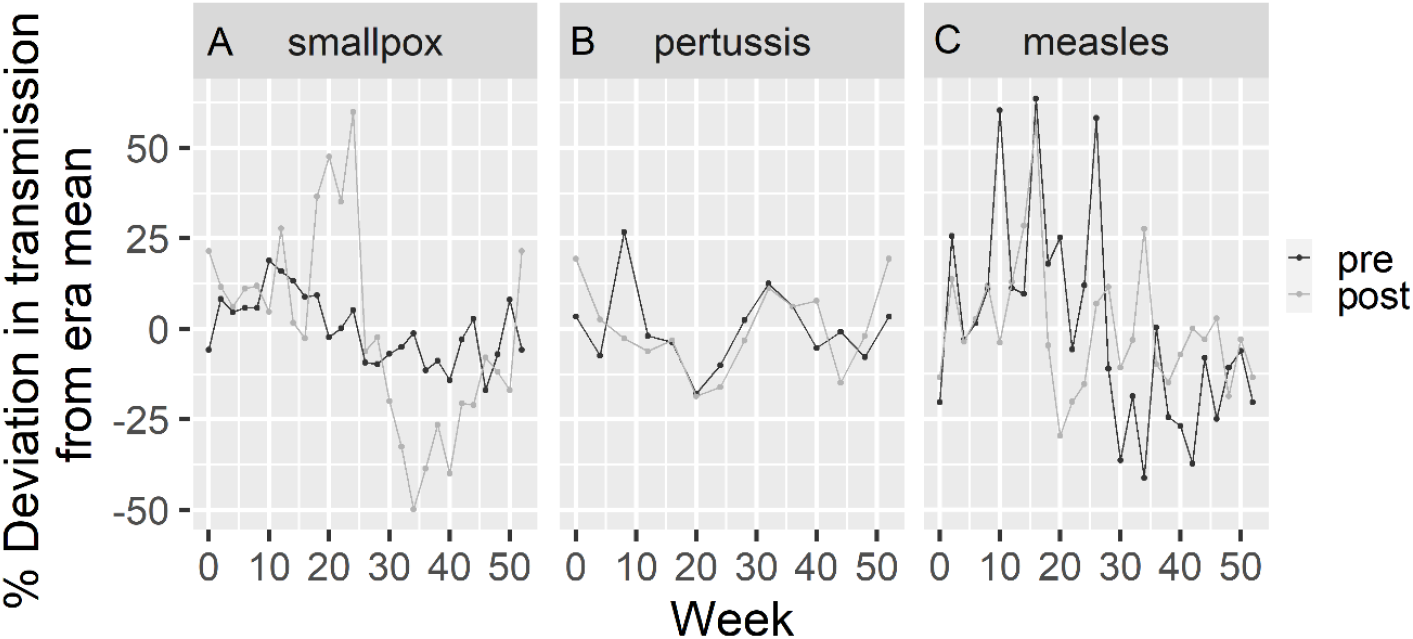
Seasonal deviation from the era’s mean transmission for (A) smallpox, (B) pertussis and (C) measles.‘Pre’ and‘post’ indicate respectively before and after the start of the smallpox vaccination programme in 1802. Note that for pertussis and measles there was no vaccine during our study period. See Fig. S1 for untransformed transmission values with 95% CI.

The seasonality of pertussis was different than that of the two other infections. In both eras, transmission increased around New Year, in late March/April and in August/September (Fig. 2B, Fig. S1B). Some of the seasonal maxima were larger than the 95% CI around the eras’ mean transmission (Fig. S1B), indicating statistically significant seasonality and this was confirmed through wavelet analyses (Fig. S2B). Hence, all three infections were seasonal, but the seasonality of pertussis was different from that of smallpox or measles.

### 3.2 Aim 2: Is the seasonality of smallpox, pertussis and measles driven by human social gathering?

Next, we tested the hypothesis that the transmission of childhood infections was higher during weeks of seasonal social gathering. In our study population, we identified five periods with seasonal social gathering events (grey weeks in Fig. 3), for a total of 32 out of 52 weeks (62%) having recurring social gathering events. These data require statistical testing for at least three reasons. First, we separated the dataset in two eras, and while this may be justified from a public health perspective, it remains unclear whether the introduction of the smallpox vaccine and the changing demography of the growing population affected the seasonality of childhood infections. Second, differently from school-term studies, where schooling occurs in relatively standardized conditions, the social gathering events in this study differed for example in their importance, group size, whether they were celebrated indoor vs. outdoor and some might increase transmission more than others. Third, seasonal social gatherings can correlate with seasonal changes in climate and we should control to what extent social events are confounded with seasonal changes in climate. To address these challenges, we estimated tSIR-based transmission rates in four-year intervals and tested statistically whether transmission rates were higher during social gathering events using GAMs.

**Fig. 3.**
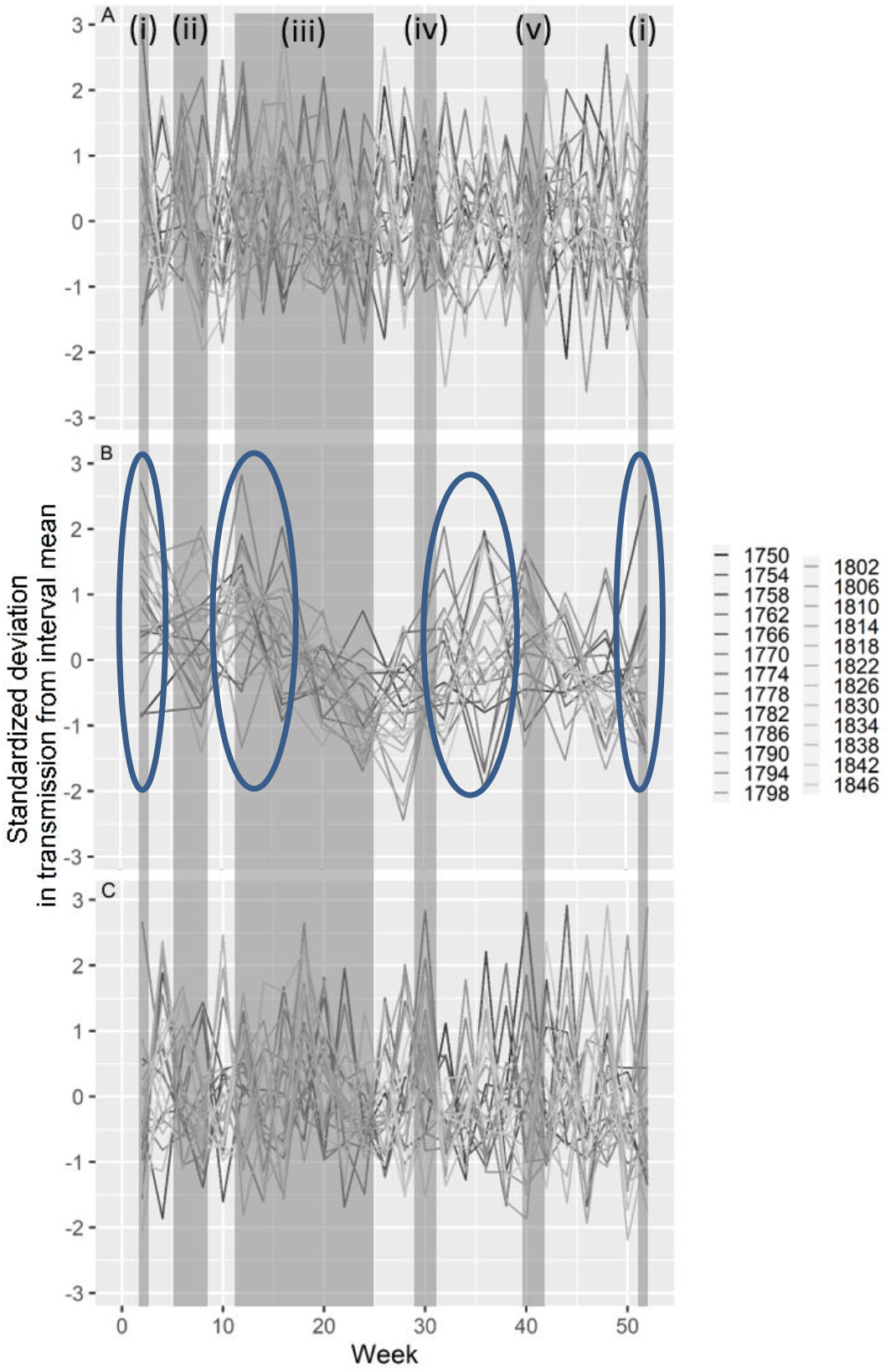
Seasonality of transmission rates estimated from tSIR models for (A) smallpox, (B) pertussis and (C) measles, showing increases in transmission during New Year in April for pertussis, but not for smallpox or measles. Shown on the Y-axis are standardized transmission coefficients (i.e. (value-mean)/sd) per four-year interval. Different lines represent four-year intervals ranging from 1750 (black) until 1850 (light grey). Dark grey zones indicate weeks of annually recurring seasonal social gathering events with (i) Christmas and New Year, (week 1 and 52), (ii) Midwinter (‘laskiainen’; week 5 to 9), (iii) the spring gatherings of Easter, Vappu, Pentecoste and Midsummer (weeks 12 to 25), (iv) summer harvest (week 28 to 32) and (v) autumn harvest (week 39 to 44). In (B) blue circles indicate periods with statistically significant increases in seasonality

The results did not support increased transmission of smallpox and measles during weeks with social gathering events (smallpox: ΔAICc=+9.6, p=0.85, model 37 in Table S1, ΔAICc=+9.4, p=0.82, Table S2; measles: ΔAICc=+4.7, p=0.09, model 57 in Table S1; ΔAICc=+1.8, p=0.13, Table S2). Furthermore, support for a gradual seasonal cycle was inconclusive at best, i.e. models with the daylight term were within 4 AICc of models without and the p-values for the daylight term in all these models were >0.05 (smallpox: ΔAICc=+2.2, p=0.96, model 21 in Table S1, ΔAICc=+1.8, p=0.12 in Table S2; measles: ΔAICc=-1.9, p=0.12, model 53 in Table S1, ΔAICc=+2.0, p=0.20 in Table S2). For neither infection did we find statistical support for era-specific seasonality (ΔAICc=+2.0, p=0.99, Table S2A & C), era-specific social gathering effects (ΔAICc>+10.1, p>0.54, Table S2A & C) or era-specific daylight effects (ΔAICc=+4.0, p>0.34, Table S2A & C).

In contrast, pertussis showed increased transmission at the end of December and early January, i.e. New Year (weeks 52 and 1, Fig. 3B), in late March and April (weeks 12 to 16, coinciding with Easter, Fig. 3B) and in August and September (weeks 32 to 40, Fig. 3B). The New Year maximum occurred as an abrupt rather than gradual event, i.e. on top of the smooth term (multivariate: ΔAICc=-18.7, p=0.0046, model 53 in Table S1; univariate: ΔAICc=-7.4, p=0.01, Table S2; Fig. 3B, Fig. S3A), but the latter two transmission periods were better described by a gradual cycle (i.e. a smooth term) rather than an abrupt event (multivariate: ΔAICc=-114.8, p<2e^-16^, model 53 in Table S1; univariate: ΔAICc=-23.3, p<e^-8^, Table S2; Fig. S3B & C). The gradual seasonal cycle was at its lowest during early summer and early winter (Fig. S3C) and hence does not show the summer-winter gradient we would expect from a uniquely climate driven seasonal phenomenon. The winter minimum in the smooth term did not cause the statistically significant increase in transmission during New Year as this increase remained statistically significant in a model without seasonal cycle (multivariate: ΔAICc=-11.8, p=0.0017, models 63 vs. 64 in Table S1; univariate: ΔAICc=-8.8, p=0.004 in Table S2B). We found no statistical support for era-specific seasonality (ΔAICc=+34.0, p=0.97, Table S2B), era-specific social gathering effects (ΔAICc=+35.1, p=0.94, Table S2B) or era-specific daylight effects (ΔAICc=+37.1, p=0.01, Table S2B). Thus, pertussis showed three seasonal maxima during New Year, in late March/April and in August/September, and the New Year maximum was abrupt, while the March/April and August/September maxima were gradual.

## 4. Discussion

In this study we quantified the seasonality of three childhood infections in a pre-industrial society without schools, 18^th^ and 19^th^ century Finland. For all three infections, we detected statistically significant seasonality. For smallpox and measles, transmission was higher in the first half of the year than in the second half, but we found no evidence that this seasonality was associated with social gathering events. In contrast, pertussis transmission increased at (i) the end of December and early January, (ii) in March and April and (iii) in August and September. The first two maxima were associated with the social gathering events New Year and Easter. Here we discuss the implications of our results for the seasonality and dynamics of infectious diseases in general and of childhood infections in particular.

A large body of work on the seasonality of measles in pre-vaccine Europe and in contemporary industrialized societies found higher transmission in winter, spring and autumn and minima during spring and summer breaks, indicating that seasonality is driven by school term-forcing (Bjørnstad et al. 2002; Metcalf et al. 2009; Mahmud et al. 2017; Klinkenberg et al. 2018). Our results show that two hundred years earlier, the seasonality of measles was very different from that in contemporary industrialized European societies with school terms. Measles showed increased transmission in the first half of the year without association with seasonal social gathering events. We believe that the lack of association with social gathering events is because Finland in the 18^th^ and 19^th^ century can best be described as a meta-population, with different regions of Finland regularly being disconnected from one another and from neighboring countries. In this meta-population, infections would often experience local or even nation-wide extinctions (Fig. 1), only to be re-introduced from other regions in Finland or from abroad. In such a scenario, the seasonality of measles could be driven by larger-scale human movement, with the increased early-year transmission (Fig. 2) being driven by the start of human movement after harsh winters during which susceptibles accumulated. Similar larger-scale seasonal human movements driving the seasonality of measles have been described, for example, in Niger, a contemporary largely agricultural based society (Ferrari et al. 2010; Bharti et al. 2011). We suggest that the seasonal dynamics of measles in our population could reflect a similar phenomenon.

We know almost nothing on the seasonality of smallpox epidemics (Krylova and Earn 2020). A recent study on the seasonality of smallpox mortality in London indicated a weak seasonal trend from summer in the 1700’s that gradually moved towards winter in the 1800’s (Krylova and Earn 2020). Our results quite differ from those in London in the same period, with smallpox transmission being predominantly in the first half of the year, a seasonality consistent with that of measles (Fig. 2). Interestingly, in the 18^th^ and early 19^th^ smallpox in London was endemic (Duncan et al. 1994; Krylova and Earn 2020). This is different from the situation in the Finnish meta-population and we believe that the seasonality of smallpox is driven by the same dynamics as that of measles, namely early-year introductions from neighboring regions through large-scale human movement.

In contrast to the predictable seasonality of measles, the seasonal dynamics of pertussis in contemporary industrialized societies are more stochastic, with a seasonality that can be age-specific and does not always match periods of school-term aggregation well (Farizo et al. 1992; Rohani et al. 2002; Skowronski et al. 2002; Metcalf et al. 2009; Domenech de Cellès et al. 2018). In North America there appears to be a dominant seasonality during August, September or in winter (Skowronski et al. 2002; Fisman et al. 2011). In England, seasonality moved from July or August in the 1940’s and 1950’s to October and December in the 1960’s and 1970’s (Fine and Clarkson 1986). Our results are different from these earlier studies in that we found increased transmission around New years and in late March/April, which matched the New Year and Easter social gatherings respectively. However, we also found a maximum from August until October (Fig. 3) and this is consistent with many previous studies, even in contemporary societies with a relatively high vaccination coverage (Farizo et al. 1992; Skowronski et al. 2002; Metcalf et al. 2009; Domenech de Cellès et al. 2018). In our study, the timing of that maximum did not match any seasonal social gathering events, which occurred either before or after the maximum in transmission (Fig. 3) and even though that seasonal maximum remains as yet unexplained (Kilgore et al. 2016; Martinez 2018), our result considerably broadens the consistency of this seasonal pattern across time and societies, supporting a possible association with some climatic factors such as humidity (Kilgore et al. 2016).

It remains puzzling as to why local social gathering events did not drive the seasonality of measles and smallpox, but did affect the seasonality of pertussis. The difference between these childhood infections may be caused by several factors and here we bring forward two potential mechanisms. First, the degree of immunization differs between infections, with immunity to pertussis wanes over time and hence transmission occurring in adults as well (Wendelboe et al. 2005), while smallpox and measles being life-long immunizing (Hammarlund et al. 2003; Griffin et al. 2012). Because of the waning immunity, pertussis in pre-vaccine eras can often transmitted by adults (Bisgard et al. 2004), while the other two infections are essentially transmitted by children (Anderson and May 1991). Such a different age-specificity can drive the seasonal differences between infections, with for pertussis social gathering events of adults being more determinant than that of children. A second noteworthy component is that pertussis is the most transmissible of the three infections. Both pertussis and measles have a high R_0_, the number of secondary infections deriving from one infected person (Anderson and May 1991), of 16 and 14, respectively (Keeling and Rohani 2011; Guerra et al. 2017). However, the R_0_ values are high for very different reasons. For pertussis, the high value arises from a high transmission probability per contact. Measles, however, has a generation time half that of pertussis and a lower transmission probability per contact. Smallpox, with an R_0_ of around 6 (Gani and Leach 2001) and a generation time of two weeks, is the infection with the lowest transmission probability per contact of the three infections studied here.

There are at least three limitations to our study. First, the causes of death in historical records were registered by parish priests, who may or not have the know-how to identify infections. To this end, we focused our analyses on three easily identifiable causes of death. Second, individuals might be more likely to die from infections in some seasons rather than others, irrespective of infection incidence. In the Northern hemisphere, the general pattern is that mortality gradually increases in winter (Rau 2007). The seasonal infection maxima in our study did not match that pattern and hence are unlikely to be driven by general seasonality in mortality. To what extent there is a possible discrepancy between the seasonality of incidence and mortality in our historical data remains yet unclear, but we are gathering historical incidence data to investigate the question. Secondly, there can be seasonal variation in access to health care or registration of information on infectious diseases. We do not believe this to be the case here, as health care was virtually non-existent and because deaths were recorded by the parish priests, who were locally-based. However, even though we believe that seasonal variation in registration of information was limited, it remains possible that for example parish priests were postponing recording information when they were busy with other activities, such as New Year or Easter. Interestingly though, for a subset of the data the time lag between death date and burial date during these two gathering events did not differ from other periods of the year (results not shown).

To conclude, here we provide a quantification of the seasonality of three childhood infections smallpox, pertussis and measles in a society for which data are rarely available, an 18^th^ and 19^th^ century pre-industrial European nation. Our results show that the seasonality of infections was very different from that in 20^th^ century industrial societies and emphasize a pathogen-specific role of local social movements as a driver of infectious disease dynamics.

## Data Availability

The data will be made available after acceptance of the manuscript

## Acknowledgements

The data are the result of years of work by the Genealogical Society of Finland, who kindly provided access to the data. We thank Jess Metcalf, Alexander Becker and Angi Rösch and the Nordemics consortium for useful discussions.

## Supplementary information

### Supplementary Information S1: Infectious disease seasonality per vaccine era based on tSIR models

**Fig. S1.**
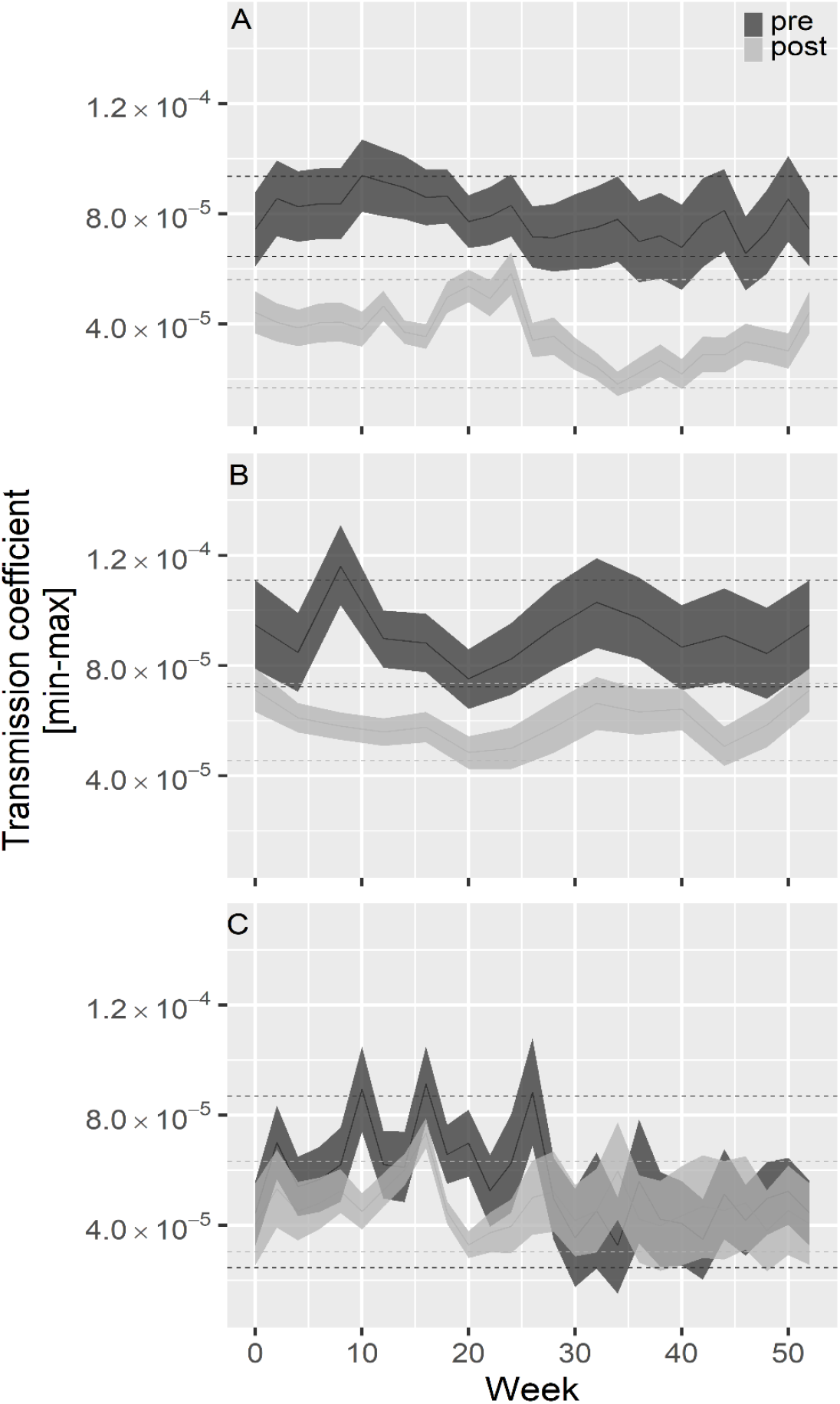
Seasonality of transmission per era estimated from tSIR models for (A) smallpox, (B) pertussis and (C) measles. Dark grey and light grey lines indicate mean, minimum and maximum transmission rates for the pre-smallpox-vaccine and the smallpox-vaccine eras respectively. Horizontal lines show 95% CI for the pre-smallpox-vaccine (dark grey) and the smallpox-vaccine (light grey) eras.

### Supplementary Information S2: Infectious disease seasonality based on wavelet analyses

**Fig S2.**
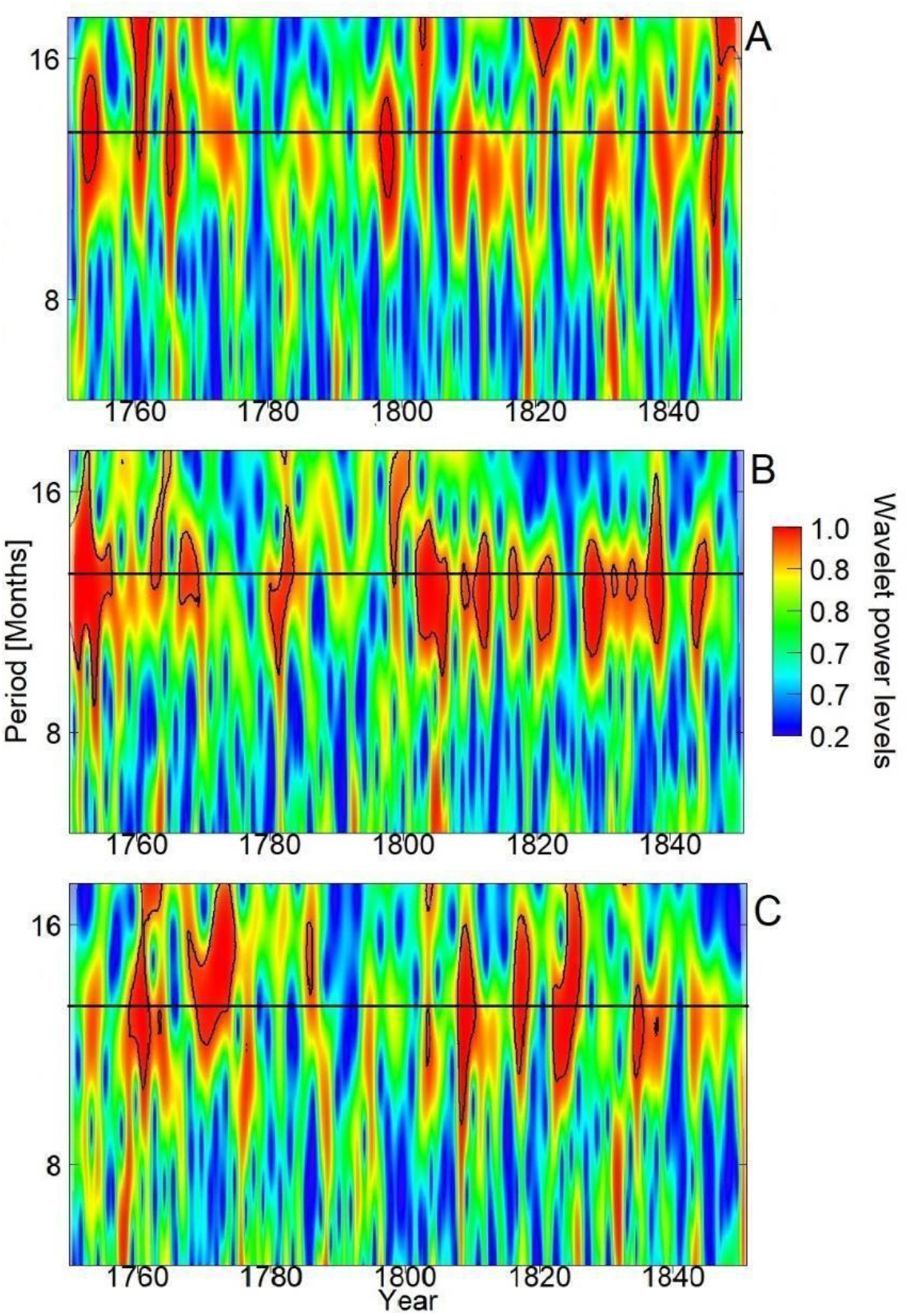
Wavelet analyses of seasonality of (A) smallpox, (B) pertussis and (C) measles between 1750 and 1850. Legends depict wavelet power levels with red referring to dominant periodicity and green or blue showing low levels of periodicity. Horizontal black lines denote the period of one year. Red zones surrounded by black lines indicate statistically significant periodicity at the 5% level.

### Supplementary Information S3: Model selection in Fig. 3

**Table S1.**
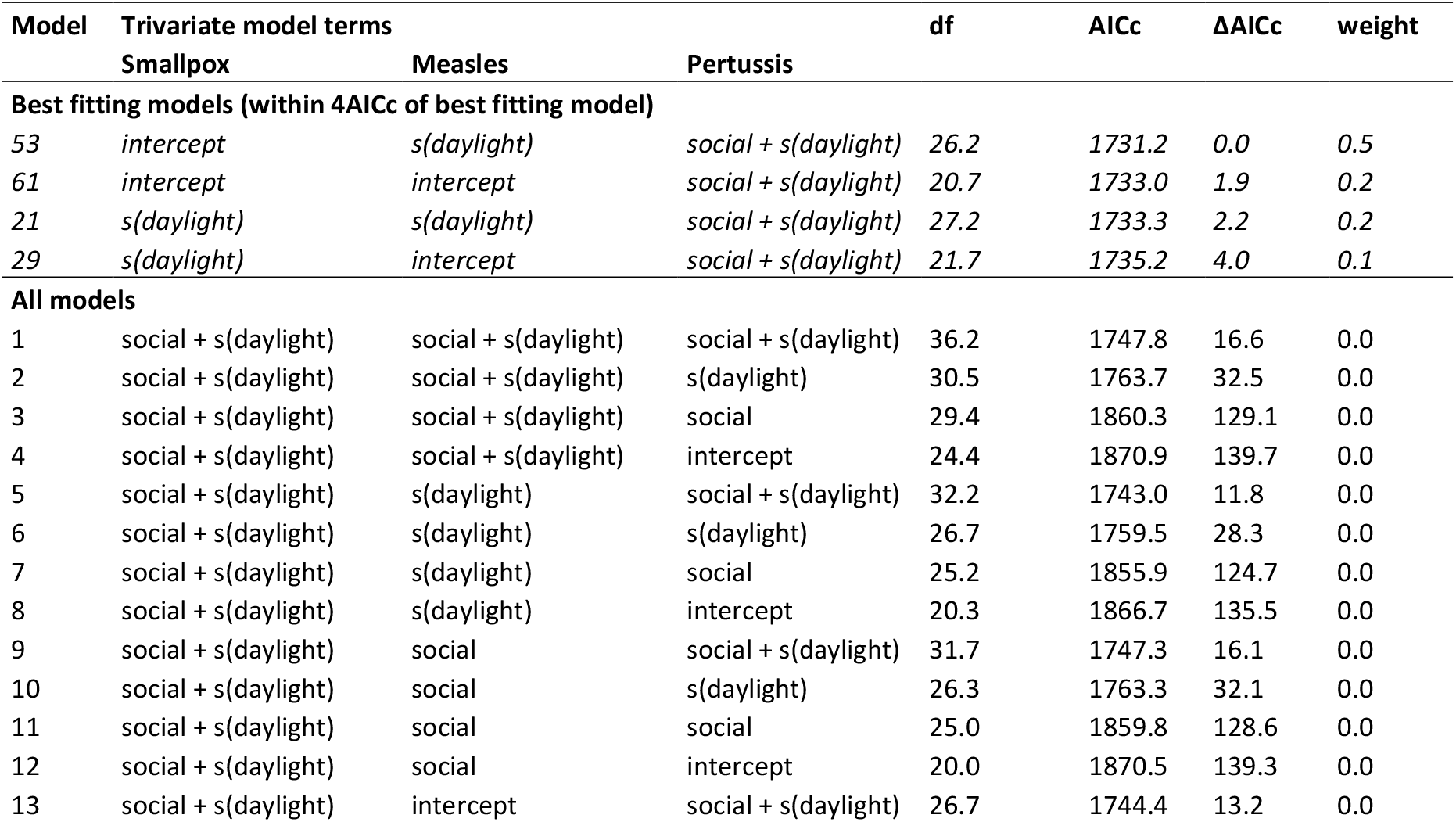
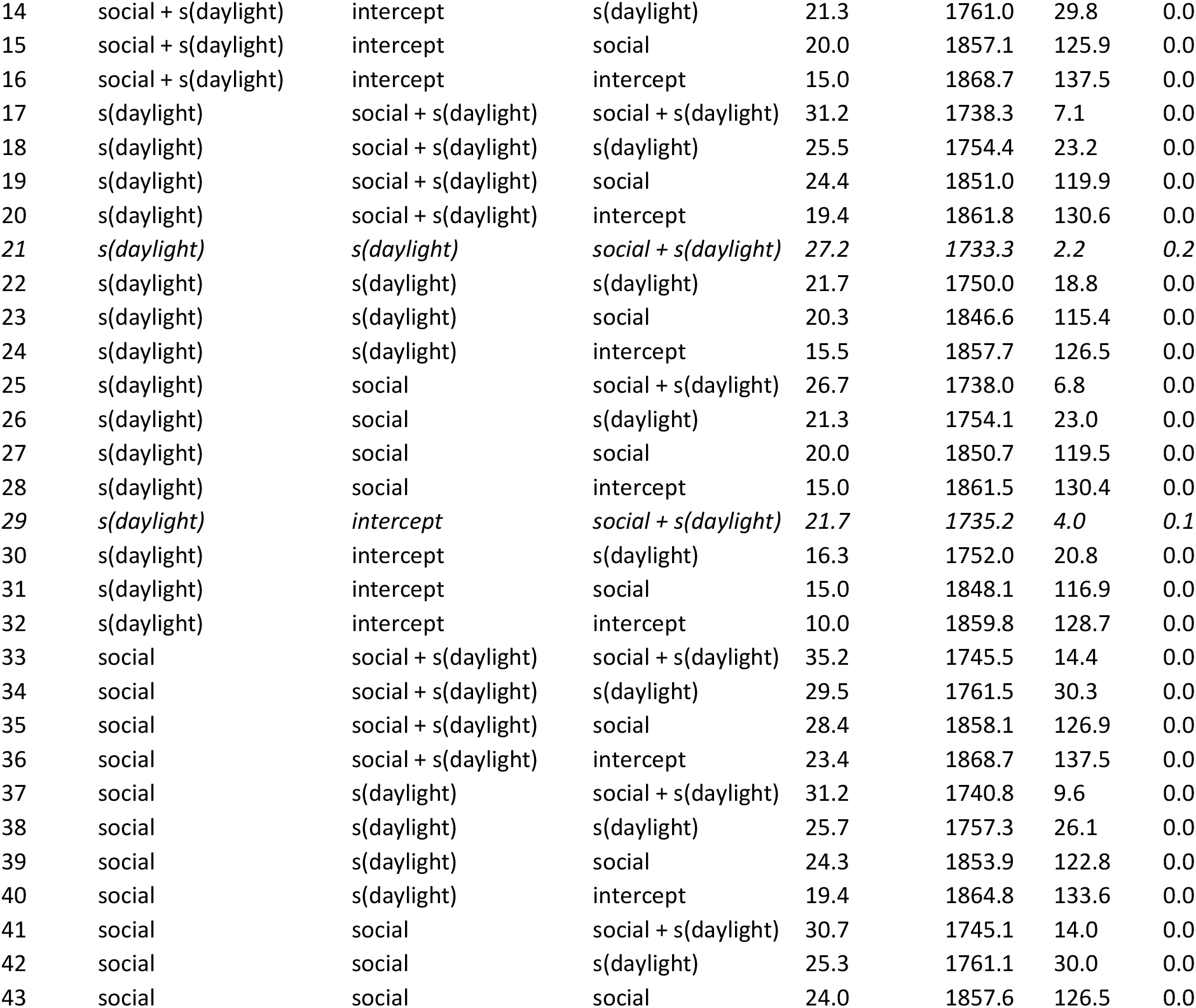
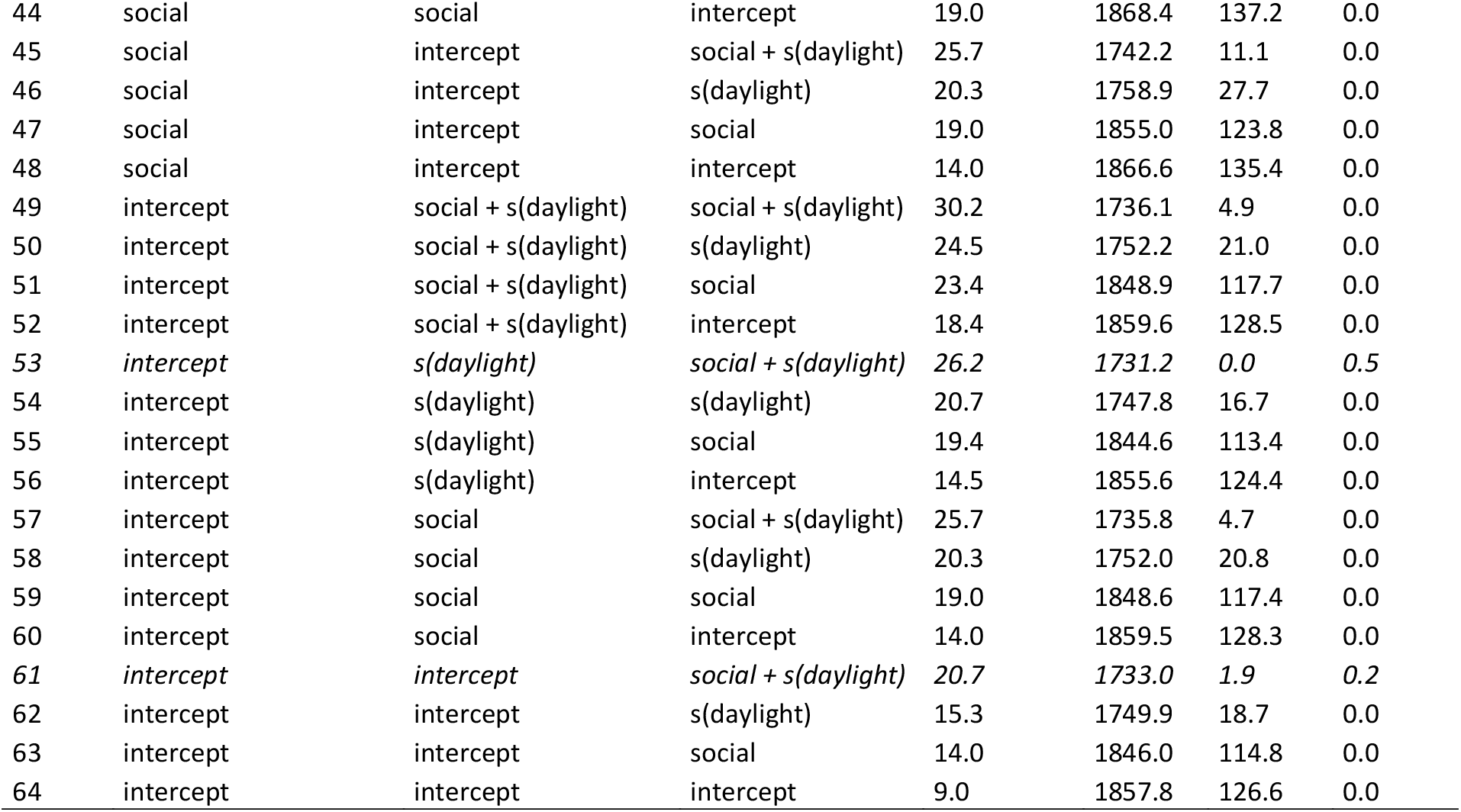
Model selection of tri-variate general additive model of tSIR-based transmission rates for the three infections. Predictor variable‘social’ captures whether or not week belonged to one of five seasonal social gathering events (six-level factor for five social gathering events and asocial weeks). To account for a possible climatic seasonal dynamic in the transmission of infection we included daylight as a cubic smoothed term shown in the table as s(daylight). Co-variation between infection transmission rates remained low, in the best fitting model (model 53): smallpox-measles: 0.094, pertussis-measles: 0.033 and smallpox-pertussis: 0.0025.

| Model | Trivariate model terms | | | df | AICc | $\Delta\text{AICc}$ | weight |
| --- | --- | --- | --- | --- | --- | --- | --- |
|  | Smallpox | Measles | Pertussis |  |  |  |  |
| **Best fitting models (within 4AICc of best fitting model)** |  |  |  |  |  |  |  |
| 53 | *intercept* | *s(daylight)* | *social + s(daylight)* | 26.2 | 1731.2 | 0.0 | 0.5 |
| 61 | *intercept* | *intercept* | *social + s(daylight)* | 20.7 | 1733.0 | 1.9 | 0.2 |
| 21 | *s(daylight)* | *s(daylight)* | *social + s(daylight)* | 27.2 | 1733.3 | 2.2 | 0.2 |
| 29 | *s(daylight)* | *intercept* | *social + s(daylight)* | 21.7 | 1735.2 | 4.0 | 0.1 |
| **All models** |  |  |  |  |  |  |  |
| 1 | *social + s(daylight)* | *social + s(daylight)* | *social + s(daylight)* | 36.2 | 1747.8 | 16.6 | 0.0 |
| 2 | *social + s(daylight)* | *social + s(daylight)* | *s(daylight)* | 30.5 | 1763.7 | 32.5 | 0.0 |
| 3 | *social + s(daylight)* | *social + s(daylight)* | *social* | 29.4 | 1860.3 | 129.1 | 0.0 |
| 4 | *social + s(daylight)* | *social + s(daylight)* | *intercept* | 24.4 | 1870.9 | 139.7 | 0.0 |
| 5 | *social + s(daylight)* | *s(daylight)* | *social + s(daylight)* | 32.2 | 1743.0 | 11.8 | 0.0 |
| 6 | *social + s(daylight)* | *s(daylight)* | *s(daylight)* | 26.7 | 1759.5 | 28.3 | 0.0 |
| 7 | *social + s(daylight)* | *s(daylight)* | *social* | 25.2 | 1855.9 | 124.7 | 0.0 |
| 8 | *social + s(daylight)* | *s(daylight)* | *intercept* | 20.3 | 1866.7 | 135.5 | 0.0 |
| 9 | *social + s(daylight)* | *social* | *social + s(daylight)* | 31.7 | 1747.3 | 16.1 | 0.0 |
| 10 | *social + s(daylight)* | *social* | *s(daylight)* | 26.3 | 1763.3 | 32.1 | 0.0 |
| 11 | *social + s(daylight)* | *social* | *social* | 25.0 | 1859.8 | 128.6 | 0.0 |
| 12 | *social + s(daylight)* | *social* | *intercept* | 20.0 | 1870.5 | 139.3 | 0.0 |
| 13 | *social + s(daylight)* | *intercept* | *social + s(daylight)* | 26.7 | 1744.4 | 13.2 | 0.0 |
| 14 | social + s(daylight) | intercept | s(daylight) | 21.3 | 1761.0 | 29.8 | 0.0 |
| 15 | social + s(daylight) | intercept | social | 20.0 | 1857.1 | 125.9 | 0.0 |
| 16 | social + s(daylight) | intercept | intercept | 15.0 | 1868.7 | 137.5 | 0.0 |
| 17 | s(daylight) | social + s(daylight) | social + s(daylight) | 31.2 | 1738.3 | 7.1 | 0.0 |
| 18 | s(daylight) | social + s(daylight) | s(daylight) | 25.5 | 1754.4 | 23.2 | 0.0 |
| 19 | s(daylight) | social + s(daylight) | social | 24.4 | 1851.0 | 119.9 | 0.0 |
| 20 | s(daylight) | social + s(daylight) | intercept | 19.4 | 1861.8 | 130.6 | 0.0 |
| 21 | *s(daylight)* | *s(daylight)* | *social + s(daylight)* | 27.2 | 1733.3 | 2.2 | 0.2 |
| 22 | s(daylight) | s(daylight) | s(daylight) | 21.7 | 1750.0 | 18.8 | 0.0 |
| 23 | s(daylight) | s(daylight) | social | 20.3 | 1846.6 | 115.4 | 0.0 |
| 24 | s(daylight) | s(daylight) | intercept | 15.5 | 1857.7 | 126.5 | 0.0 |
| 25 | s(daylight) | social | social + s(daylight) | 26.7 | 1738.0 | 6.8 | 0.0 |
| 26 | s(daylight) | social | s(daylight) | 21.3 | 1754.1 | 23.0 | 0.0 |
| 27 | s(daylight) | social | social | 20.0 | 1850.7 | 119.5 | 0.0 |
| 28 | s(daylight) | social | intercept | 15.0 | 1861.5 | 130.4 | 0.0 |
| 29 | *s(daylight)* | *intercept* | *social + s(daylight)* | 21.7 | 1735.2 | 4.0 | 0.1 |
| 30 | s(daylight) | intercept | s(daylight) | 16.3 | 1752.0 | 20.8 | 0.0 |
| 31 | s(daylight) | intercept | social | 15.0 | 1848.1 | 116.9 | 0.0 |
| 32 | s(daylight) | intercept | intercept | 10.0 | 1859.8 | 128.7 | 0.0 |
| 33 | social | social + s(daylight) | social + s(daylight) | 35.2 | 1745.5 | 14.4 | 0.0 |
| 34 | social | social + s(daylight) | s(daylight) | 29.5 | 1761.5 | 30.3 | 0.0 |
| 35 | social | social + s(daylight) | social | 28.4 | 1858.1 | 126.9 | 0.0 |
| 36 | social | social + s(daylight) | intercept | 23.4 | 1868.7 | 137.5 | 0.0 |
| 37 | social | s(daylight) | social + s(daylight) | 31.2 | 1740.8 | 9.6 | 0.0 |
| 38 | social | s(daylight) | s(daylight) | 25.7 | 1757.3 | 26.1 | 0.0 |
| 39 | social | s(daylight) | social | 24.3 | 1853.9 | 122.8 | 0.0 |
| 40 | social | s(daylight) | intercept | 19.4 | 1864.8 | 133.6 | 0.0 |
| 41 | social | social | social + s(daylight) | 30.7 | 1745.1 | 14.0 | 0.0 |
| 42 | social | social | s(daylight) | 25.3 | 1761.1 | 30.0 | 0.0 |
| 43 | social | social | social | 24.0 | 1857.6 | 126.5 | 0.0 |
| 44 | social | social | intercept | 19.0 | 1868.4 | 137.2 | 0.0 |
| 45 | social | intercept | social + s(daylight) | 25.7 | 1742.2 | 11.1 | 0.0 |
| 46 | social | intercept | s(daylight) | 20.3 | 1758.9 | 27.7 | 0.0 |
| 47 | social | intercept | social | 19.0 | 1855.0 | 123.8 | 0.0 |
| 48 | social | intercept | intercept | 14.0 | 1866.6 | 135.4 | 0.0 |
| 49 | intercept | social + s(daylight) | social + s(daylight) | 30.2 | 1736.1 | 4.9 | 0.0 |
| 50 | intercept | social + s(daylight) | s(daylight) | 24.5 | 1752.2 | 21.0 | 0.0 |
| 51 | intercept | social + s(daylight) | social | 23.4 | 1848.9 | 117.7 | 0.0 |
| 52 | intercept | social + s(daylight) | intercept | 18.4 | 1859.6 | 128.5 | 0.0 |
| 53 | *intercept* | *s(daylight)* | *social + s(daylight)* | 26.2 | 1731.2 | 0.0 | 0.5 |
| 54 | intercept | s(daylight) | s(daylight) | 20.7 | 1747.8 | 16.7 | 0.0 |
| 55 | intercept | s(daylight) | social | 19.4 | 1844.6 | 113.4 | 0.0 |
| 56 | intercept | s(daylight) | intercept | 14.5 | 1855.6 | 124.4 | 0.0 |
| 57 | intercept | social | social + s(daylight) | 25.7 | 1735.8 | 4.7 | 0.0 |
| 58 | intercept | social | s(daylight) | 20.3 | 1752.0 | 20.8 | 0.0 |
| 59 | intercept | social | social | 19.0 | 1848.6 | 117.4 | 0.0 |
| 60 | intercept | social | intercept | 14.0 | 1859.5 | 128.3 | 0.0 |
| 61 | *intercept* | *intercept* | *social + s(daylight)* | 20.7 | 1733.0 | 1.9 | 0.2 |
| 62 | intercept | intercept | s(daylight) | 15.3 | 1749.9 | 18.7 | 0.0 |
| 63 | intercept | intercept | social | 14.0 | 1846.0 | 114.8 | 0.0 |
| 64 | intercept | intercept | intercept | 9.0 | 1857.8 | 126.6 | 0.0 |

**Table S2.** Model selection of the univariate general additive model of tSIR-based transmission rates for (A) smallpox, (B) pertussis and (C) measles. Predictor variable‘social’ captures whether or not week belonged to one of five seasonal social gathering events (six-level factor for five social gathering events and asocial weeks). To account for a possible climatic seasonal dynamic in the transmission of infection we included daylight as a cubic smoothed term shown in the table as s(daylight). For each infection, the best model is shown in italic.

| Model | df | AICc | $\Delta AICc$ | weight |
| --- | --- | --- | --- | --- |
| **(A) Smallpox** |  |  |  |  |
| social + s(daylight) | 10.0 | -12042.4 | 11.5 | 0.0 |
| s(daylight) | 5.0 | -12051.8 | 2.0 | 0.2 |
| social | 9.0 | -12044.4 | 9.4 | 0.0 |
| *intercept* | 4.0 | -12053.8 | 0.0 | 0.5 |
| era | 5.0 | -12051.8 | 2.0 | 0.2 |
| era*social | 15.0 | -12037.8 | 16.1 | 0.0 |
| era*s(daylight) | 6.0 | -12049.8 | 4.0 | 0.1 |
| **(B) Pertussis** |  |  |  |  |
| *social + s(daylight)* | 10.0 | -5984.0 | 0.0 | 1.0 |
| s(daylight) | 5.0 | -5976.6 | 7.4 | 0.0 |
| social | 9.0 | -5960.6 | 23.3 | 0.0 |
| intercept | 4.0 | -5951.9 | 32.1 | 0.0 |
| era | 5.0 | -5949.8 | 34.2 | 0.0 |
| era*social | 15.0 | -5948.9 | 35.1 | 0.0 |
| era*s(daylight) | 6.0 | -5946.9 | 37.1 | 0.0 |
| **(C) Measles** |  |  |  |  |
| social + s(daylight) | 10.0 | -11219.7 | 3.2 | 0.1 |
| s(daylight) | 5.0 | -11220.9 | 2.0 | 0.2 |
| social | 9.0 | -11221.1 | 1.8 | 0.2 |
| *intercept* | 4.0 | -11222.9 | 0.0 | 0.5 |
| era | 5.0 | -11220.9 | 2.0 | 0.2 |
| era*social | 15.0 | -11212.8 | 10.1 | 0.0 |
| era*s(daylight) | 6.0 | -11218.9 | 4.0 | 0.1 |

### Supplementary Information S4: GAM correlations and partial residuals of the model in Fig. 3

**Fig S3.**
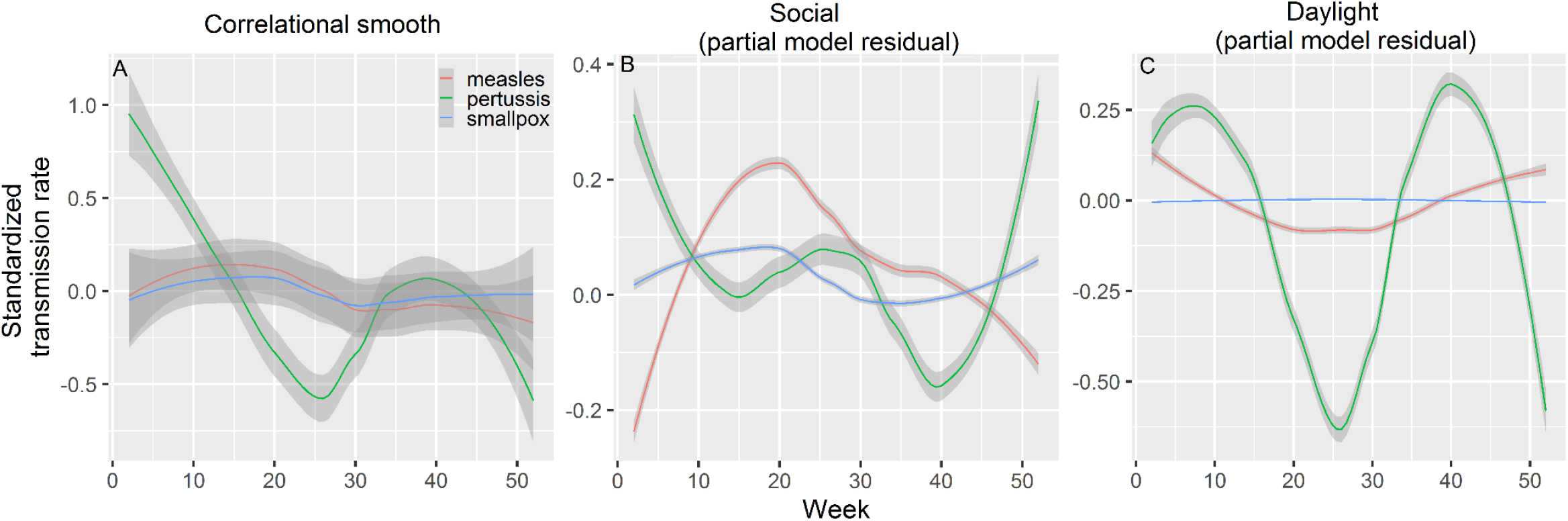
Seasonal changes in standardized transmission rates with (A) data correlation, and (B) and (C) the decomposed contributions (partial residuals) of respectively the social and daylight terms in the full trivariate model (model 1 in Table S2).

## Notes

### Competing Interest Statement

The authors have declared no competing interest.

### Funding Statement

We are grateful to the Academy of Finland (278751, 292368), Nordforsk (104910), the Ehrnrooth Foundation, the Finnish Cultural Foundation, the University of Turku Foundation and the Doctoral Programme in Biology, Geography and Geology, University of Turku.

## References

Altizer, S., A. Dobson, P. Hosseini, P. Hudson, and M. Pascual. 2006. Seasonality and the dynamics of infectious diseases. Ecology Letters 9:467–484.

Anderson, R. M., and R. May. 1991. Infcetious diseases of humans. Oxford University Press, Oxford, UK.

Bakker, K. M., M. E. Martinez-Bakker, B. Helm, and T. J. Stevenson. 2016. Digital epidemiology reveals global childhood disease seasonality and the effects of immunization. Proceedings of the National Academy of Sciences 113:6689–6694.

Becker, A. D., and B. T. Grenfell. 2017. tsiR: An R package for time-series Susceptible-Infected-Recovered models of epidemics. PloS One 12:e0185528.

Benenson, A., ed. 1981. Control of Communicable Disease in Man (13th ed.). American Public Health Association, Washington, DC.

Bharti, N., A. Tatem, M. Ferrari, R. Grais, A. Djibo, and B. T. Grenfell. 2011. Explaining seasonal fluctuations of measles in Niger using nighttime lights imagery. Science 334:1424–1428.

Bisgard, K. M., F. B. Pascual, K. R. Ehresmann, C. A. Miller, C. Cianfrini, C. E. Jennings, C. A. Rebmann, et al. 2004. Infant pertussis: Who was the source? Pediatric Infectious Disease Journal 23:985–989.

Bjørnstad, O. N., B. F. Finkelstädt, and B. T. Grenfell. 2002. Dynamics of measles epidemics: estimating scaling of transmission rates using a time series SIR model. Ecological Monographs 72:169–184.

Burnham, K. P., and D. R. Anderson. 2002. Model selection and multimodel inference: A practical information-theoretic approach (2nd ed.). Springer-Verlag, New York.

Burnham, K. P., D. R. Anderson, and K. P. Huyvaert. 2011. AIC model selection and multimodel inference in behavioral ecology: Some background, observations, and comparisons. Behavioral Ecology and Sociobiology 65:23–35.

Cauchemez, S., N. M. Ferguson, C. Wachtel, A. Tegnell, G. Saour, B. Duncan, and A. Nicoll. 2009. Closure of schools during an influenza pandemic. The Lancet Infectious Diseases 9:473–481.

Cauchemez, S., A. J. Valleron, P. Y. Boëlle, A. Flahault, and N. M. Ferguson. 2008. Estimating the impact of school closure on influenza transmission from Sentinel data. Nature 452:750–754.

Caudron, Q., A. S. Mahmud, C. J. E. Metcalf, M. Gottfreðsson, C. Viboud, A. D. Cliff, B. T. Grenfell, et al. 2015. Predictability in a highly stochastic system: final size of measles epidemics in small populations. Journal of the Royal Society Interface 12:20141125.

Cazelles, B., M. Chavez, G. Constantin de Magny, J.-F. Guégan, and S. Hales. 2007. Time-dependent spectral analysis of epidemiological time-series with wavelets. Journal of The Royal Society Interface 4:625–636.

Centers for Disease Control and Prevention. 2002. Pertussis Deaths--United States, 2000. Morbidity and Mortality Weekly Report 51:616–618.

Conlan, A. J. K., and B. T. Grenfell. 2007. Seasonality and the persistence and invasion of measles. Proceedings of the Royal Society B: Biological Sciences 274:1133–1141.

Deguen, S., G. Thomas, and N. P. Chau. 2000. Estimation of the contact rate in a seasonal SEIR model: Application to chickenpox incidence in France. Statistics in Medicine 19:1207–1216.

Domenech de Cellès, M., F. M. G. Magpantay, A. A. King, and P. Rohani. 2018. The impact of past vaccination coverage and immunity on pertussis resurgence. Science Translational Medicine 10:eaaj1748.

Downie, A. W., D. S. Fedson, L. S. Vincent, A. R. Rao, and C. H. Kempe. 1969. Haemorrhagic smallpox. Journal of Hygiene 67:619–629.

Duncan, C. J., S. R. Duncan, and S. Scott. 1996. Whooping cough epidemics in London, 1701-1812: Infection dynamics, seasonal forcing and the effects of malnutrition. Proceedings of the Royal Society B: Biological Sciences 263:445–450.

Duncan, C. J., S. R. Duncan, and S. Scott. 1997. The dynamics of measles epidemics. Theoretical Population Biology 52:155–163.

Duncan, S. R., S. Scott, and C. J. Duncan. 1994. Modelling the different smallpox epidemics in England. Philos. Trans.R.Soc.Lond B Biol.Sci. 346:407–419.

Eames, K. T. D., N. L. Tilston, E. Brooks-Pollock, and W. J. Edmunds. 2012. Measured dynamic social contact patterns explain the spread of H1N1v influenza. PLoS Computational Biology 8:e1002425.

Earn, D. J. D., P. Rohani, B. M. Bolker, and B. T. Grenfell. 2000. A simple model for complex dynamical transitions in epidemics. Science 287:667–671.

Farizo, K. M., S. L. Cochi, E. R. Zell, E. W. Brink, G. Wassilak, P. A. Patriarca, S. Clinical, et al. 1992. Epidemiological features of pertussis in the United States, 1980-1989. Clinical Infectious Diseases 14:708–719.

Ferrari, M. J., A. Djibo, R. F. Grais, N. Bharti, B. T. Grenfell, and O. N. Bjornstad. 2010. Rural – urban gradient in seasonal forcing of measles transmission in Niger. Proceedings of the Royal Society B Biological Sciences 277:2775–2782.

Ferrari, M. J., R. F. Grais, N. Bharti, A. J. K. Conlan, O. N. Bjørnstad, L. J. Wolfson, P. J. Guerin, et al. 2008. The dynamics of measles in sub-Saharan Africa. Nature 451:679–684.

Fine, P. E. M., and J. A. Clarkson. 1982. Measles in England and Wales. An analysis of factors underlying seasonal patterns. International Journal of Epidemiology 11:5–15.

Fine, P. E. M., and J. A. Clarkson. 1986. Seasonal influences on pertussis. International Journal of Epidemiology 15:237–247.

Fisman, D. N., P. Tang, T. Hauck, S. Richardson, S. J. Drews, D. E. Low, and F. Jamieson. 2011. Pertussis resurgence in Toronto, Canada: A population-based study including test-incidence feedback modeling. BMC Public Health 11.

Gani, R., and S. Leach. 2001. Transmission potential of smallpox in contemporary populations. Nature 414:748–751.

Grassly, N. C., and C. Fraser. 2006. Seasonal infectious disease epidemiology. Proceedings of the Royal Society B Biological Sciences 273:2541–2550.

Griffin, D. E., W. H. Lin, and C. H. Pan. 2012. Measles virus, immune control, and persistence. FEMS Microbiology Reviews 36:649–662.

Guerra, F. M., S. Bolotin, G. Lim, J. Heffernan, S. L. Deeks, Y. Li, and N. S. Crowcroft. 2017. The basic reproduction number (R0) of measles: a systematic review. The Lancet Infectious Diseases 17:e420–e428.

Guyer, B., and A. M. Mcbean. 1981. The epidemiology and control of measles in Yaoundé, Cameroun, 1968 - 1975. International Journal of Epidemiology 10:263–269.

Hammarlund, E., M. W. Lewis, S. G. Hansen, L. I. Strelow, J. A. Nelson, G. J. Sexton, J. M. Hanifin, et al. 2003. Duration of antiviral immunity after smallpox vaccination. Nature Medicine 9:1131– 1137.

Holopainen, J., and S. Helama. 2009. Little ice age farming in Finland: Preindustrial agriculture on the edge of the Grim reaper’s scythe. Human Ecology 37:213–225.

Itkonen, T. 1948. The lapps of Finland up to 1945, vol.2. Werner Söderström Osakeyhtiö, Porvoo, Helsinki.

Karjalainen, S. 1994. Juhlan aika: Suomalaisia vuotuisperinteitä. W Söderström, Helsinki, Finland.

Keeling, M. J., and P. Rohani. 2011. Modeling infectious diseases in humans and animals. Princeton University Press, Princeton, USA.

Ketola, T., M. Briga, T. Honkola, M. Vuotillainen, and V. Lummaa. 2021. Town population size and structuring into villages and households drive infectious disease risks in pre-healthcare Finland. Proceedings of Royal Society B Biological Sciences 288:20210356.

Kilgore, P. E., A. M. Salim, M. J. Zervos, and H. Schmitt. 2016. Pertussis: microbiology, disease, treatment, and prevention. Clinical Microbiology Reviews 29:449–486.

Klinkenberg, D., S.JM. Hahné, T. Woudenberg, and J. Wallinga. 2018. The reduction of measles transmission during school vacations. Epidemiology 29:562–570.

Krylova, O., and D. J. D. Earn. 2020. Patterns of smallpox mortality in London, England, over three centuries. PLoS Biology 18:1–27.

London, W. P., and J. A. Yorke. 1973. Recurrent outbreaks of measles, chickenpox and mumps: I. Seasonal variation in contact rates. American Journal of Epidemiology 98:453–468.

Mahmud, A. S., C. J. E. Metcalf, and B. T. Grenfell. 2017. Comparative dynamics, seasonality in transmission, and predictability of childhood infections in Mexico. Epidemiology and Infection 145:607–625.

Martinez, M. E. 2018. The calendar of epidemics: Seasonal cycles of infectious diseases. PLoS Pathogens 14:e1007327.

Metcalf, C. J. E., O. N. Bjørnstad, B. T. Grenfell, and V. Andreasen. 2009. Seasonality and comparative dynamics of six childhood infections in pre-vaccination Copenhagen. Proceedings of the Royal Society B Biological Sciences 276:4111–4118.

Metcalf, C. J. E., K. S. Walter, A. Wesolowski, C. O. Buckee, E. Shevliakova, A. J. Tatem, W. R. Boos, et al. 2017. Identifying climate drivers of infectious disease dynamics: recent advances and challenges ahead. Proceedings of Royal Society B Biological sciences 284:20170901.

Official Statistics of Finland. 2018. Available at: https://www.stat.fi/index_en.html. Accessed: November 1st.

Pitkänen, K. 2007. Suomen väestön historialliset kehityslinjat [Historical development of the Finnish population]. Pages 41–76 in S. Koskinen, T. Martelin, I.-L. Notkola, V. Notkola, K. Pitkänen, M. Jalovaara, E. Mäenpää, et al., eds. Suomen väestö [The Finnish population]. Helsinki University Press, Helsinki, Finland.

Rau, R. 2007. Seasonality in human mortality: a demographic approach. Springer-Verlag, Berlin Heidelberg.

Roesch, A., and H. Schmidbauer. 2018. WaveletComp: Computational Wavelet Analysis.

Rohani, P., C. Green, N. Mantilla-Beniers, and B. Grenfell. 2003. Ecological interference between fatal diseases. Nature 422:885–888.

Rohani, P., M. J. Keeling, and B. T. Grenfell. 2002. The interplay between determinism and stochasticity in childhood diseases. American Naturalist 159:469–481.

Saarivirta, T., D. Consoli, and P. Dhondt. 2012. The evolution of the Finnish health-care system early 19th Century and onwards. International Journal of Business and Social Science 3:243– 258.

Skowronski, D., G. De Serres, D. MacDonald, W. Wu, C. Shaw, J. Macnabb, S. Champagne, et al. 2002. The changing age and seasonal profile of pertussis in Canada. Journal of Infectious Diseases 186:1537–1538.

Stone, L., R. Olinky, and A. Huppert. 2007. Seasonal dynamics of recurrent epidemics. Nature 446:533–536.

Tiimonen, S. 2001. Valoa kansalle: Luterilainen kirkko ja kansanopetuksen kehittämispyrkimykset autonomisessa Suomessa 1809–1848. Suomen kirkkohistoriallisen seuran toimituksia 185. Suomen kirkkohistoriallinen seura, Helsinki.

Vilkuna, K. 1950. Vuotuinen ajantieto: Vanhoista merkkipäivistä sekä kansanomaisesta talous-ja sääkalenterista enteillen (25th ed.). Otava.

Voutilainen, M., J. Helske, and H. Högmander. 2020. A Bayesian Reconstruction of a Historical Population in Finland, 1647–1850. Demography 57:1171–1192.

Vuorinen, H. S. 1999. Suomalainen tautinimistö ennen bakteriologista vallankumousta. Hippokrates 33–61.

Wendelboe, A. M., A. Van Rie, S. Salmaso, and J. A. Englund. 2005. Duration of immunity against pertussis after natural infection or vaccination. Pediatric Infectious Disease Journal 24:58–61.

Woods, S. 2017. Generalized Additive Models. An Introduction with R (Second Ed.). Chapman & Hall, Boca Raton, Fl.

Woods, S., N. Pya, and B. Safken. 2016. Smoothing parameter and model selection for general smooth models (with discussion). Journal of the American Statistical Association 111:1548– 1575.

